# The effect of GLP-1RA exenatide on Idiopathic Intracranial Hypertension: Randomised Clinical Trial

**DOI:** 10.1101/2022.05.24.22275518

**Authors:** James L. Mitchell, Hannah S. Lyons, Jessica K. Walker, Andreas Yiangou, Olivia Grech, Zerin Alimajstorovic, Nigel H. Greig, Yazhou Li, Georgios Tsermoulas, Kristian Brock, Susan P. Mollan, Alexandra J. Sinclair

## Abstract

Therapeutics to reduce intracranial pressure are an unmet need. Pre-clinical data has demonstrated a novel strategy to lower intracranial pressure using glucagon-like peptide-1 receptor signaling.

Here, we translate these findings into patients by conducting a randomized, placebo controlled, double-blind trial to assess the effect of exenatide, a glucagon-like peptide-1 receptor agonist, on intracranial pressure in idiopathic intracranial hypertension. Telemetric intracranial pressure catheters enabled long-term intracranial pressure monitoring. The trial enrolled adult women with active idiopathic intracranial hypertension (intracranial pressure >25 cmCSF and papilloedema) who receive subcutaneous exenatide or placebo. The three primary outcome measures were intracranial pressure at 2.5 hours, 24 hours and 12 weeks and alpha set a priori at less than 0.1.

Among the 16 women recruited, 15 completed the study (mean age 28 ± 9, body mass index 38.1 ± 6.2 kg/m^2^, intracranial pressure 30.6 ± 5.1 cmCSF). Exenatide significantly and meaningfully lowered intracranial pressure at 2.5 hours -5.7 ± 2.9 cmCSF (*p=0*.*048*); 24 hours of -6.4 ± 2.9 cmCSF (*p=0*.*030*); and 12 weeks -5.6 ± 3.0 cmCSF (*p=0*.*058*). No serious safety signals were noted.

This data provides confidence to proceed to a phase 3 trial in idiopathic intracranial hypertension and highlights the potential to utilise glucagon-like peptide-1 receptor agonist in other conditions characterised by raised intracranial pressure.

## Introduction

Glucagon-like peptide-1 (GLP-1) is a gut neuropeptide secreted by the distal small intestine in response to a meal.^1^ GLP-1 receptor agonists are existing therapeutic agents used to in the treatment of diabetes. GLP-1 stimulates glucose-dependant insulin secretion and inhibits glucagon release, thereby lowering blood glucose but not causing hypoglycaemia. ^2^ GLP-1 receptor agonists also signal at the hypothalamus to regulate satiety and weight.^3^ This has led to GLP-1 receptor agonists being licensed to promote weight loss in the setting of obesity.^4^

Relevant to this trial is the role of GLP-1 receptor agonists in regulating fluid secretion. GLP-1 receptor agonists have been shown to reduce sodium reabsorption and promote diuresis through actions in the renal proximal tubule.^5,6^ GLP-1 receptors are expressed in the choroid plexus, the predominant cerebrospinal fluid secreting structure in the brain.^7,8^ Our preliminary data has shown that GLP-1 receptor agonism reduces CSF secretion and intracranial pressure (ICP) in an *in vivo* rodent model with elevated ICP.^7^ The reduction in ICP was of a greater magnitude to that observed with the commonly used drugs in idiopathic intracranial hypertension.^9^

Idiopathic Intracranial Hypertension is characterized by increased intracranial pressure ICP with no identifiable cause. Recent weight gain is the major risk factor for development of the condition and its occurrence is most commonly observed in women of reproductive age with obesity.^10,11^ Weight loss is disease modifying and if maintained can induce remission.^12,13^ As the incidence of idiopathic intracranial hypertension is rising^14-16^ in line with global obesity trends,^17^ targeted treatments are an unmet clinical need.

Visual loss is observed in greater than 90% of those with idiopathic intracranial hypertension^18^ with up to 25% suffering severe permanent blindness.^19^ Chronic disabling headaches occur in the majority and have adverse impact on quality of life.^20,21^ Cognitive deficits linked to raised ICP have been documented.^22^ Currently, there is no licenced therapy for idiopathic intracranial hypertension.^23^ The most commonly used off-label medicine is acetazolamide, but due to side effects and treatment failures new therapies are needed. Patient groups have highlighted the importance of prioritising novel targeted treatments for idiopathic intracranial hypertension.^24^

The aim of this study was to translate the pre-clinical data demonstrating efficacy of GLP-1 receptor signalling to reduce ICP into patients with raised ICP by conducting a randomized, placebo controlled, double-blind trial in idiopathic intracranial hypertension. The trial aimed to evaluate both acute effects on ICP as well as effects over a 3-month time horizon.

## Materials and methods

### Trial Design and Oversight

The trial was a prospective, randomised, parallel group, placebo-controlled trial in women with active idiopathic intracranial hypertension. Patients with a diagnosis of active idiopathic intracranial hypertension were identified and recruited from a single tertiary referral hospital (University Hospitals Birmingham NHS Foundation Trust). This study was approved by the West Midlands - Solihull Research Ethics Committee (17/WM/0179) and all subjects provided written informed consent according to Declaration of Helsinki principles. The trial was registered with ISTCRN (12678718).

### Participants

Women aged 18-60 years who met the diagnostic criteria for idiopathic intracranial hypertension were recruited.^25^ All had normal brain imaging, including magnetic resonance venography or computed tomography venography (apart from radiological signs of raised ICP). All eligible patients had active idiopathic intracranial hypertension (optic nerve head swelling in at least one eye and ICP >25cmCSF). Those with significant co-morbidities, prior CSF diversion procedures, those currently using GLP-1 receptor agonists or DPP-4 inhibitors or taking drugs that were thought to reduce ICP were excluded. Those taking drugs that might influence ICP discontinued these at least a month prior to enrolment. Pregnant patients or those planning pregnancy were excluded, urine HCG was checked at each study visit. Detailed enrolment criteria are provided in the Supplementary Table 1.

### Assessments

Following enrolment, a 28-day headache diary was completed (capturing monthly headache days (MHD), headache severity (0-10 numerical rating sale (NRS)), monthly analgesia days). A telemetric intracranial pressure catheter (Raumedic, Germany) was implanted prior to the baseline visit.

At baseline, medical history, examination (including blood pressure and heart rate) and body mass index (BMI) (calculated using the formula BMI = (weight (kg) / height (m)^2^) were recorded and a urine pregnancy test performed. Visual assessments included LogMAR visual acuity and intraocular pressure as measured with the iCare IC200 (Main-line, UK); perimetric mean deviation (PMD) using the Humphrey visual field analyser (24-2 Swedish Interactive Threshold Algorithm (SITA) standard test pattern using a size III white stimulus). Papilloedema was confirmed on a dilated slit lamp examination by a neuro-ophthalmologist. It was quantified by spectral domain optical coherence tomography (OCT; Spectralis, Heidelberg Engineering, Germany) using the global peripapillary retinal nerve fibre layer. Patient reported outcome measures were assessed (Headache Impact Test -6 (HIT-6)^26^ and 36-item short from survey (SF-36) Rand version).^27^ Assessments were repeated at 12 weeks. Drug compliance was monitored based on the remaining exenatide in the pen injector device.

Blood samples were collected at baseline pre-dose and post-dose at 2.5, 6, 11, 22 and 24 hours, at 2 weeks (pre and 2.5 hours post-dose), and 12 weeks (pre and 2.5 hours post-dose), to evaluate safety blood tests, pharmacokinetics and antidrug antibodies.

ICP was recorded using a transdermal telemetric ICP monitoring system (Raumedic, Germany) at baseline, 2.5 hours, 24 hours and 12 weeks. In the planned exploratory analysis ICP was measured continuously for the first 2.5 hours after dosing and then between 0000hrs and 0700hours overnight following the first dose. ICP data was collected at a frequency of 5Hz and the mean ICP was calculated from each 30 minutes of continuous ICP monitoring over the first 2.5hrs and each hour overnight. The schedule of assessments is detailed in Supplementary Table 2.

### Body composition

Dual energy x-ray absorptiometry (DEXA) was performed using a total-body scanner (QDR 4500; Hologic, Bedford, MA, USA), as previously described^28,29^ on a subset of patients. The scans were conducted by a clinical scientist and trained radiographer. Patients with metal prosthetics or implants were included, and tissue overlying the prosthesis was excluded from analysis. Scans were checked for accuracy of fields of measurement. Regional fat mass was analysed as described previously.^28,29^ The precision of total fat mass measures in terms of coefficients of variation (CV) was less than 3%, and for regional fat analyses it was less than 5%. All subjects were analysed on the same DEXA scanner.

### Randomization and Study Treatment

Participants were randomized in a 1:1 ratio to either active treatment with exenatide (Byetta) or placebo using a computer-generated randomization list generated by the Birmingham Clinical Trials Unit. Treatment allocation was blinded to patient and investigators. A double check of allocation was performed by an unblinded nurse and pharmacist. The first dose was a loading dose of subcutaneous exenatide 20mcg or equivalent volume of subcutaneous 0.9% saline placebo. Subjects were then dosed for 12 weeks (self-administered at home) with either subcutaneous exenatide 10mcg or equivalent volume of placebo twice daily.

### Blood analysis

The following fasted blood tests were processed at the hospital laboratory: (creatinine (μmol/L), alanine aminotransferase (ALT) (IU/L), high-density lipoproteins (HDL) (mmol/L), total cholesterol (mmol/L), triglycerides (TG) (mmol/L), haemoglobin A1c (HbA1C) (mmol/mol)). Samples not analysed immediately were centrifuged (10 minutes at 1500 g at 4°C) aliquoted and stored at -80 °C. All samples only underwent a single freeze-thaw cycle.

### Fasting insulin and HOMA2-IR

Fasting insulin (Mercodia, Uppsala, Sweden) was measured using a commercially available assay, according to the manufacturer’s instructions. Homeostasis model assessment of insulin resistance (HOMA2-IR) was calculated using the program HOMA calculator v2.2.3. ^30^

### Pharmacokinetics

Exenatide concentration was evaluated by ELISA. ELISA was performed on serum samples from 7 patients receiving the active drug. Serum was collected at 10 time points; baseline, 2.5 hours, 6 hours, 11 hours, 22 hours and 24 hours, week 2 at baseline and 2.5 hours post dose, week 12 at baseline and 2.5 post dose. An exenatide fluorescent ELISA (Phoenix Pharmaceuticals Inc) was used. This was a competitive enzyme immunoassay, wherein the primary antibody is competitively bound by either a biotinylated peptide or the targeted peptide in samples. All samples were run in triplicate. The assay was performed according to manufacturer’s protocol.^31^

### Anti-Exendin-4 antibody levels in serum

In the light of reports of the development of anti-exenatide antibody in human studies administering exenatide in type 2 diabetes mellitus^32,33^ (with anti-exenatide titres peaking between 6-22 weeks), as well as their presence in preclinical studies,^31,34^ we investigated whether such antibodies developed in our clinical study. As the trial lasted for a period of 12 weeks, we evaluated serum anti-exendin-4 antibody levels in 16 patients at baseline (prior to exenatide treatment), and at week 2 and 12 of the trial by employing a previously developed sandwich ELISA.^34^ All week 2 and 12 serum samples were obtained before exenatide administration for the day, and serial dilutions of each serum samples were used in the ELISA. A brief protocol for the ELISA is as following: plates were coated with exenatide (exendin-4) (2 µg/ml concentration in coating buffer (AnaSpec Inc., Fremont, CA, USA)) at 4°C overnight. Thereafter, following blocking and washing steps, standards (mouse monoclonal anti-exendin-4 antibody (Abcam (ab23407), Waltham, MA, USA) and unknown samples with serial dilutions were added to the plate and incubated at room temperature for 1 hr. After washing, biotinylated-exendin-4 (2 µg/ml concentration (AnaSpec Inc., Fremont, CA, USA)) was added and followed by washing and SA-HRP detection (KPL, Gaithersburg, MD, USA). The titres of the anti-exenatide antibody within the samples were then estimated by serial dilution of the serum (to a maximum dilution of 1:125).

### Outcomes Measure

The primary outcome was ICP at 2.5hrs, 24hrs and 12 weeks post drug administration. ICP was recorded with p-Tel telemetric intracranial pressure catheter and MPR-1 reader (Raumedic, Germany). ICP was recorded continuously for 30-minute periods at specified timepoints in a standardised supine position. For the first 2.5 hours post dosing, mean ICP was calculated from each 30 minutes of continuous ICP monitoring. For the overnight recording the mean ICP was calculated from each hour of continuous ICP monitoring. ICP was sampled at 5Hz. Recordings were downloaded and analysed in Dataview version 1.2 (Raumedic, Germany). ICP was recorded in mmHg (conversion factor to cmCSF was 1.36).

Secondary outcomes included: monthly headache days, headache severity and monthly analgesia days, logarithm of the minimum angle of resolution (logMAR) visual acuity (VA) measured using the Early Treatment Diabetic Retinopathy Study (ETDRS) charts; perimetric mean deviation (PMD) using Humphrey 24-2 Swedish Interactive Thresholding Algorithm (SITA) central threshold automated perimetry; BMI and health-related quality of life (measured by short-form (SF)-36 and HIT-6). Evaluations were at baseline 2.5 hours, 24 hours and 12 weeks (with additional blood sampling at 2 weeks).

### Adverse Event Reporting

Adverse events were recorded as was drug compliance (unused medication in the injector pens documented).

### Sample Size Calculation

In a study of 25 patients, Sinclair et al.^13^ showed that the cross-sectional sample standard deviation of ICP is 4.9 – 5.1 cmCSF, measured at baseline and immediately before and after a longitudinal intervention (low energy diet). There are very few trials in idiopathic intracranial hypertension and the minimal clinically important change for LP pressure is not established and may vary with individual patients. Seeking significance at least *alpha < 0*.*1* and power at least 80% using equal group sizes, a total sample size of 14 patients was required, i.e. seven patients randomised to receive active treatment and a further seven to receive control. This calculation assumed an effect size of 6.5 cmCSF with a standard deviation of 5.1 cmCSF, the upper end of the range observed previously. Allowing for 10% drop-out, the proposed recruitment was eight patients per arm, and 16 patients in total.

### Statistical Analyses

All primary analyses (primary and secondary outcomes including safety outcomes) were evaluated by intention-to-treat (ITT) analysis. Analysis was completed on received data, with every effort made to follow-up participants to minimize potential for bias. Final analyses were conducted after the final visit of the final patient of the main trial once the data had been cleaned and locked; then unblinded. No imputation of missing data was conducted. The analysis of visual data included data from the most affected eye at baseline as defined by PMD, analysis of intra-ocular pressure was performed on the mean average of both eyes. Statistical analysis was performed in R v4.0.0 (R Foundation for Statistical Computing, Vienna, Austria). Data were reported as means and SD (with median and interquartile range [IQR] for non-normal data), and SE and 95% confidence intervals (CI) where appropriate. Hierarchical linear regression models were used to analyse repeated measures of the primary and secondary outcomes and to estimate differences adjusted for baseline values. In these models, population-level effects (also known as fixed effects) comprised the intercept, time as a factor variable, and the two-way interaction of treatment arm and time as a factor variable to model changing treatment effects over time. Group-level effects (also known as random effects) comprised patient-level adjustments to the intercept. The threshold for statistical significance was pre-specified at 0.1.

### Data availability

Anonymized individual participant data will be made available along with the trial protocol and statistical analysis plan. Proposals should be made to the corresponding author and will be reviewed by the Data Sharing Committee in discussion with the Chief Investigator. A formal Data Sharing Agreement may be required between respective organizations once release of the data is approved and before data can be released.

## Results

### Patients

Between November 1st, 2017, and September 17th, 2018, 18 participants were screened, 16 enrolled and 15 randomly assigned to either the exenatide group (*n=7*) or the placebo group (*n=8*) (Figure 1).

**Figure 1.**
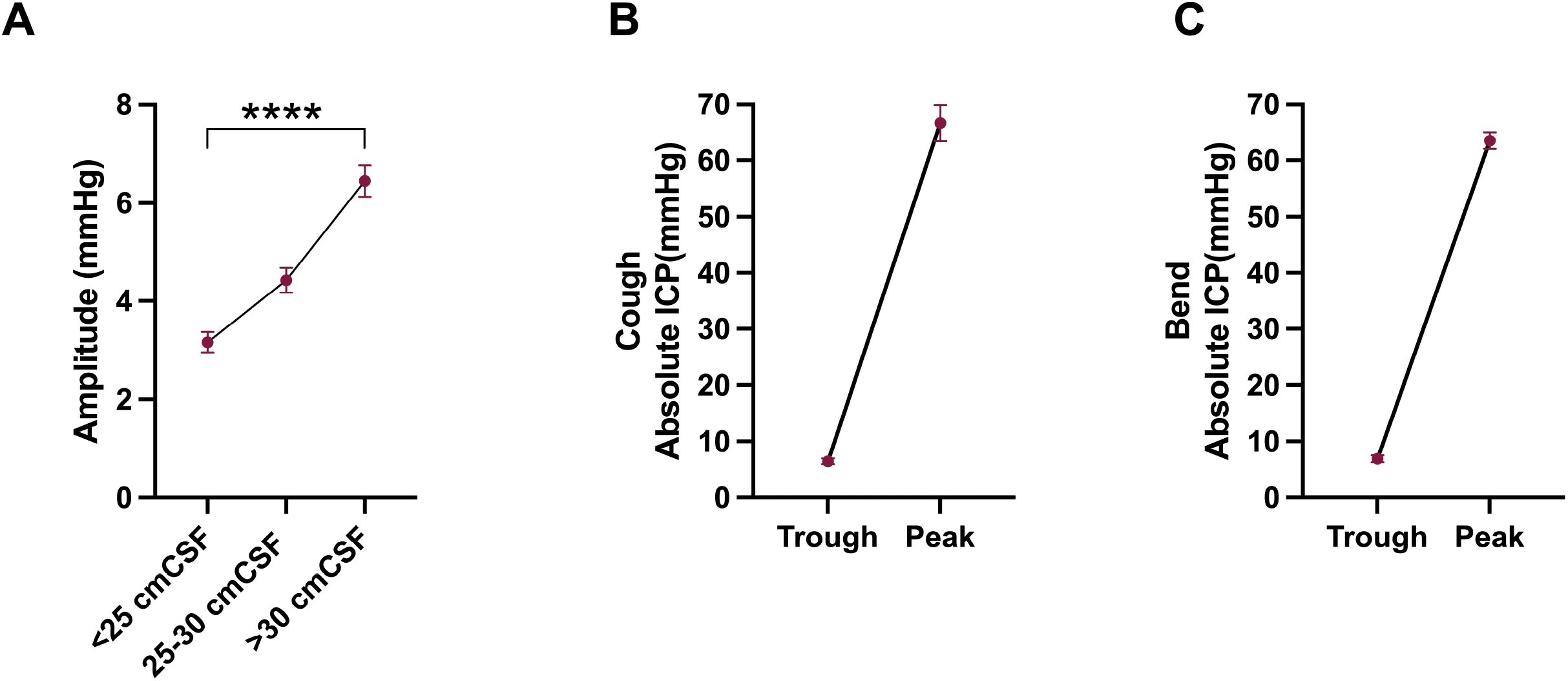
Consort diagram. Consort diagram describing the numbers and disposition of study subjects.

### Adherence to the Protocol

One participant was withdrawn before randomization due to disease progression requiring urgent cerebrospinal fluid shunting (Figure 1). All randomised patients completed the 12-week duration of the trial, however one patient experienced telemeter failure prior to week 12, therefore ICP data was not recorded, and all other data for week 12 was collected for this patient. There was full adherence to protocol with no cross over and no significant protocol deviation, except blood tests for pharmacokinetics which were missed on four occasions.

### Demographics and baseline characteristics

Baseline characteristics confined a cohort of patients with active idiopathic intracranial hypertension (Supplementary Table 3). Age, body mass index (BMI) and ICP at baseline were well-matched between groups. Median (IQ range) time from surgical implantation of ICP monitor to baseline visit was 10 days (16.5) (Supplementary Table 3). At baseline there was a significant difference in monthly headache days between arms, exenatide 21.6 (5.2) (mean (SD)) and placebo 10.3 (8.5); there was also a significant difference in perimetric mean deviation, exenatide -0.6 (1.0)dB (mean (SD) and placebo -2.7 (1.9) dB (Supplementary Table 4).

### Primary outcome measure

The primary clinical outcome was the difference in ICP between exenatide and placebo, as measured by telemetric ICP monitoring at 2.5 hours, 24 hours and 12 weeks. The difference in ICP between arms at 2.5 hours was -4.2 (2.1) (mean (SD) mmHg), *p=0*.*048*, and at 24 hours was -4.7 (2.1), *p=0*.*030*. The effect was sustained at 12 weeks with an ICP difference of -4.1 (2.2), *p=0*.*058*. (Table 1, Figure 2) This was equivalent to 5.7 (2.9) cmCSF (Mean (SD)) at 2.5 hours, 6.4 (2.9) cmCSF at 24 hours and 5.6 (3.9) cmCSF at 12 weeks.

**Figure 2.**
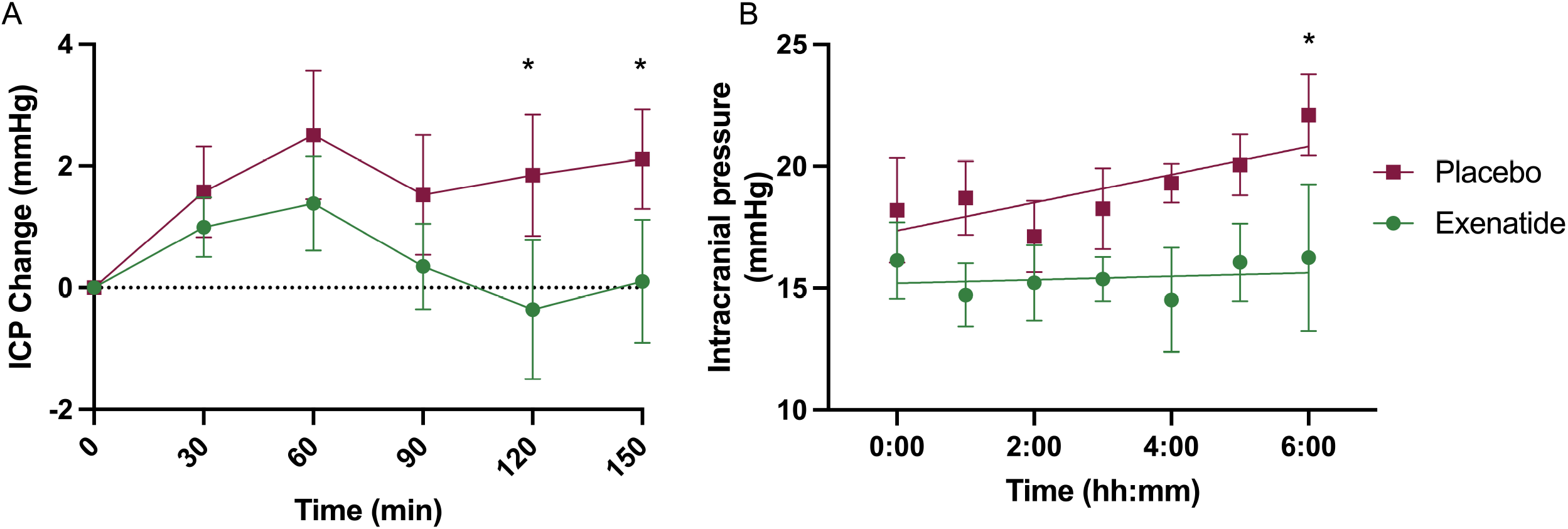
Primary outcomes. Mean change (SEM) of ICP in (**A**) ICP at 2.5 hours; (**B**) ICP at 24 hours ; (**C**) ICP at 12 weeks. \**P<0*.*1* (significant level set at *P<0*.*1*). Diurnal variability in ICP reflected in changes in ICP readings at different times of day observed in the placebo arm.

### Secondary clinical outcomes

The key secondary outcome was monthly headache days which reduced significantly in the exenatide arm, -7.7 (9.2) days (mean (SD), *p=0*.*069* compared to the placebo arm -1.5 (4.8) days, *p=0*.*404* (Figure 3), although there was no significant difference between arms at 12 weeks. Changes in monthly analgesia days showed a trend to improvement in the exenatide arm, but not in the placebo arm (Figure 3, Supplementary Table 4). There was no significant change in headache severity (Supplementary Table 5). Headache disability measured by HIT-6 was significantly higher in the exenatide arm at baseline 62.9 (3.2) vs 55.8 (6.9), (mean (SD)), *p=0*.*041*, there was no significant change over the course of the trial (Figure 3, Supplementary Table 6).

**Figure 3.**
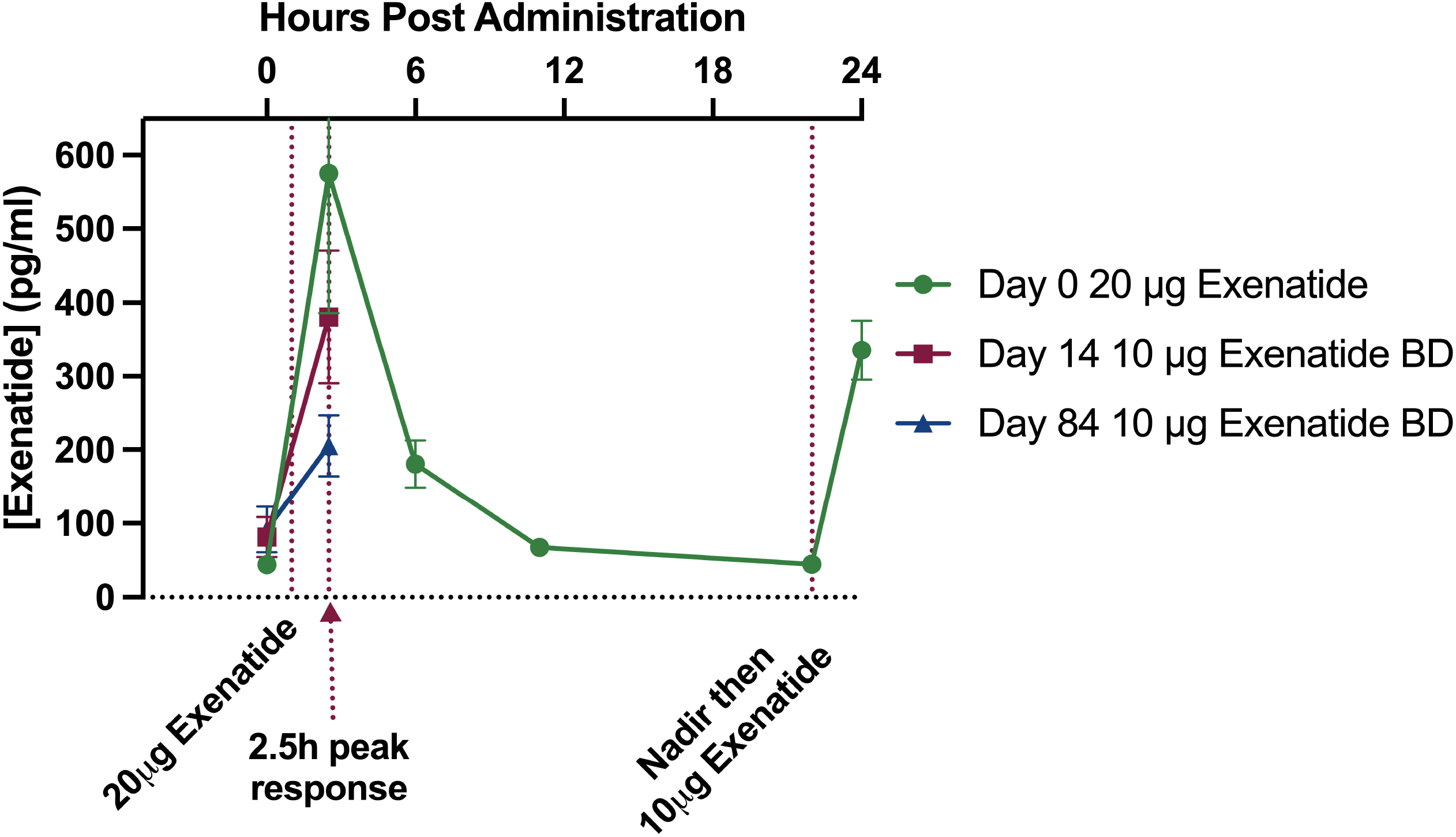
Monthly headache days and analgesia, Visual acuity and BMI. Mean change (SEM) at 12 weeks of (**A**) Monthly headache days, (**B**) Monthly analgesia days, (**C**) Visual acuity measured by logMAR and (**D**) Body mass index (BMI). \**P<0*.*1* (significant level set at *P<0*.*1*).

### Vision

Visual acuity significantly improved in the exenatide arm compared to the placebo arm -0.1 (0.05) LogMar units, *p=0*.*036*. There was no significant change in perimetric mean deviation, difference between arms 1.0 (0.8) dB, *p=0*.*188*. Optical Coherence Tomography Retinal Nerve Fibre Layer (OCT RNFL) did not change significantly between arms, -40.2 (47.2) µm, *p=0*.*396*. (Figure 3, Supplementary Table 4).

### Intra-ocular pressure

There was no significant change in intra-ocular pressure, difference between arms -0.1 (1.1), *p=0*.*910* (Supplementary Table 4).

### Quality of life

Analysis of quality of life, utilizing the SF36, showed no significant changes in either the physical or mental component scores (Supplementary Table 4).

### Body Mass Index

BMI in the placebo arm baseline was 38.6 (4.7) kg/m^2^ and 38.1(4.9) kg/m^2^ at 12 weeks, whereas in the exenatide arm baseline was 37.6 (7.9) kg/m^2^ and 37.5 (7.4) kg/m^2^ at 12 weeks. There was no significant difference in BMI between arms at 12 weeks, *p=0*.*854* (Figure 3, Supplementary Table 4).

### Safety blood test results

No significant changes were seen in the safety monitoring blood tests (Supplementary Table 7).

### Adverse Events

12 adverse events were reported during the trial. Eight adverse events occurred in the exenatide arm, seven of which were nausea related to exenatide initiation. Four adverse events occurred in the placebo arm. One unrelated serious adverse event, thyrotoxicosis, was reported in the placebo arm. No patients withdrew due to adverse events (Supplementary Table 8). All participants remained compliant with assigned treatment as monitored by returned medication.

### Anti-exenatide Antibodies

No anti-exenatide antibodies were detected in samples collected at baseline. In total two of the seven patients on exenatide developed antibodies by week 12. At week 2 one patient had antibodies present (> 0.1 µg/ml), these were also present at 12 weeks (> 0.1 µg/ml). A further patient was noted to have antibodies at 12 weeks (>1.0 µg/ml).

### Drug concentrations measurements

The 2 patients with anti-exenatide antibodies at 12 weeks had higher (3- and 6-fold higher) than predicted exenatide concentrations at 12 weeks and were excluded from the 12 weeks data analysis. At baseline, week 2 and week 12, there was a sharp increase in exenatide concentrations in patient serum at 2.5 hours following subcutaneous administration of exenatide. This was followed by a sharp decline in peptide concentrations at 6 hours. The mean exenatide concentrations at 2.5 hours are higher at baseline than at week 2 and week 12 (575.4, 380.7 and 205.4 pg/ml respectively), which is expected as 20 μg of exenatide was administered at baseline, compared to 10 μg administered at the week 2 and 12 (Supplementary Table 9, Supplementary Figure 1). The number of participants was insufficient for meaningful analysis of the relationship between pharmacokinetics and efficacy.

### Glucose and Insulin

No hypoglycaemia was encountered during the trial. There was no significant difference in fasted glucose and fasted insulin between the two trial arms at 12 weeks (Supplementary Table 10, Supplementary Figure 2).

### Vital Signs

Blood pressure and heart rate were stable during the trial, mean arterial pressure was lower at 12 weeks in the exenatide group compared to placebo, -6.7mmHg (3.6), *p=0*.*088*, but there was no significant change within arms between baseline and 12 weeks (Supplementary Table 4).

### Post Hoc Analysis - Exploratory ICP

We looked in more detail at the changes in ICP in the first 2.5 hours following drug administration. ICP was significantly lower recordings over time points 90-120 minutes (-4.6 (2.1) (mean (SD)) mmHg *p=0*.*028*) and 120-150 minutes (-4.2 (2.1) (mean (SD)) mmHg, *p=0*.*042*) (Supplementary Table 11, Figure 4A).

**Figure 4.**
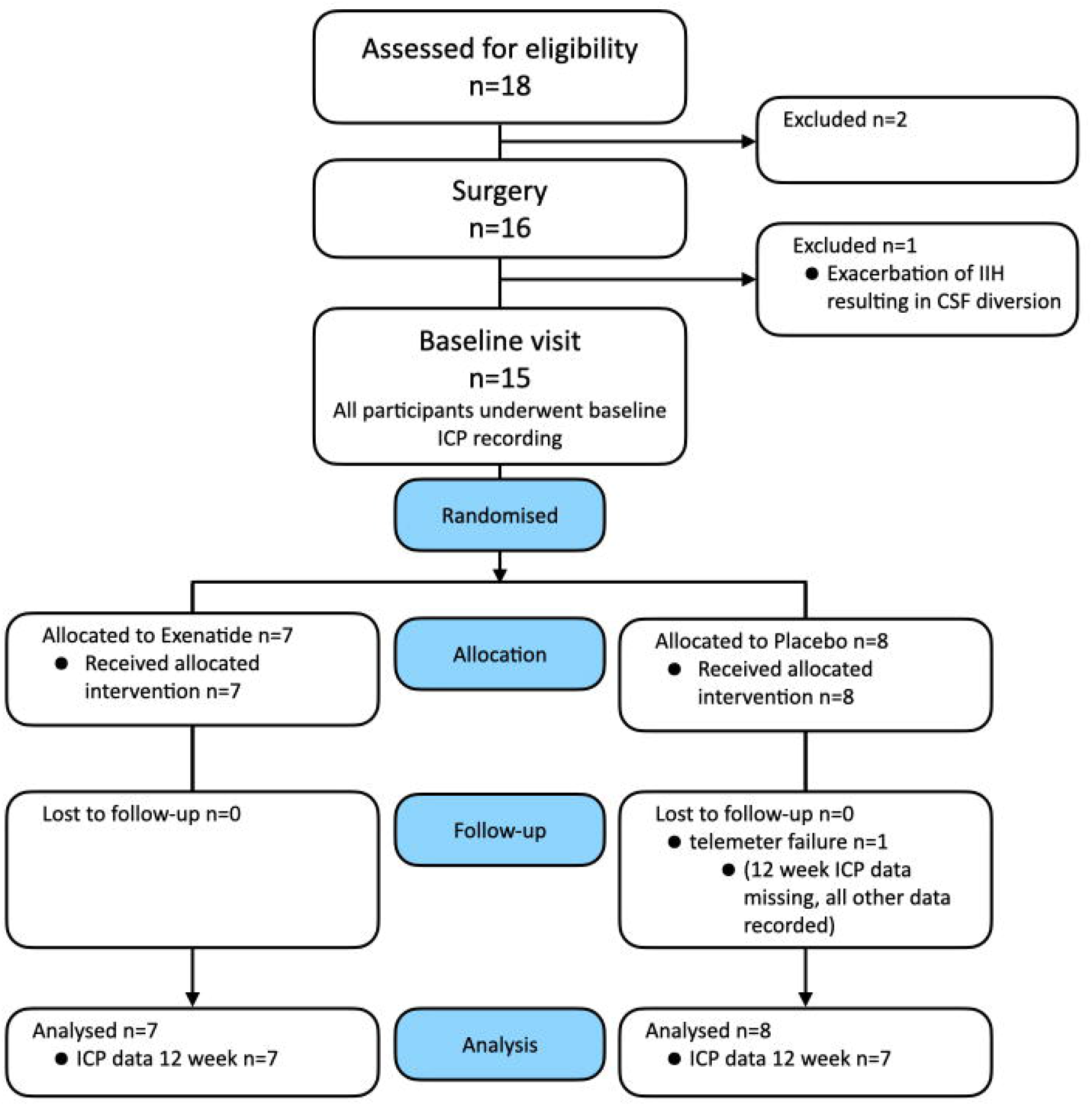
First 2.5hrs, overnight ICP. Mean ICP (SEM) by arm measured (**A**) continuously over 2.5hrs after dose and (**B**) hourly overnight. \**P<0*.*1* (significant level set at *P<0*.*1*).

ICP was then measured overnight between 2400 hours and 0700 hours on the first day of dosing (Supplementary Table 12, Figure 4B). ICP is observed to rise progressively overnight in idiopathic intracranial hypertension.^35^ In the placebo arm the ICP rose overnight as expected by 3.1 (4.1) mmHg, *p=0*.*13*. In the exenatide arm, the overnight rise in ICP was supressed to 0.1 (3.2) mmHg, *p=0*.*97*. There was a significant difference in the ICP measurement between the exenatide and placebo arm at the last reading of the night (0600-0700 hrs, ICP difference between arms was 5.0 (2.5) mmHg (mean (SD)) (equivalent to 6.8 (3.4) cm CSF), *p=0*.*04*).

### Dual energy x-ray absorptiometry

We determined that idiopathic intracranial hypertension patients had a similar fat mass, lean mass and android-gynoid ratio in those treated with exenatide as compared to those on placebo with no significant differences detected at 12 weeks (Supplementary Table 13).

## Discussion

IIH:Pressure is the first randomised controlled trial comparing the efficacy of the GLP-1 receptor agonist exenatide, with placebo to reduce ICP in patients with active idiopathic intracranial hypertension. We have found a significant and clinically meaningful reduction in ICP in the treatment arm as compared to the placebo arm both acutely and after 12 weeks of dosing. The drug was safe and well-tolerated. This trial establishes the successful translation of pre-clinical data into humans and is the first to demonstrate a new drug treatment for idiopathic intracranial hypertension that significantly reduces ICP in idiopathic intracranial hypertension.

The hallmark of idiopathic intracranial hypertension is raised ICP which causes headaches and visual loss (through compression of the optic nerve and papilloedema).^10,21,36^ Therefore, to evaluate the efficacy of exenatide the primary outcome for this trial was chosen as ICP.^37^ A significant reduction in ICP was seen acutely at 2.5 hours of -4.2mmHg (equivalent to -5.7 cmCSF), which corresponds to peak exenatide serum levels. The reduction in ICP was also noted at 24 hours (reduction of 4.7 mmHg (equivalent to 6.4 cmCSF)) (Table 1, Figure 2). Other drugs (used off-label) to treat idiopathic intracranial hypertension have not been shown to reduce ICP within the first 24 hours of administration in patients. Idiopathic intracranial hypertension patients can deteriorate rapidly (over days)^38^ and hence a drug with rapid onset of action is clinically advantageous for idiopathic intracranial hypertension. A drug treatment that can rapidly reduce ICP is also of potential relevance for other conditions characterised by raised ICP such as following traumatic brain injury and stroke.

A significant reduction in ICP was also noted after prolonged dosing at 12 weeks (-4.1 mmHg equivalent to -5.6 cmCSF) (Table 1, Figure 2), demonstrating durability of dosing. The minimal clinically important change in ICP in those with idiopathic intracranial hypertension has yet to be determined.^37^ There have been four previous randomised control trials in idiopathic intracranial hypertension^12,39-41^ and one prospective crossover trial.^13^ Four of these have utilised ICP as an outcome measure, with the magnitude of treatment effect between trial arms ranging from 5.9 cmCSF (acetazolamide and diet);^40^ −6.0 (1.8) (by bariatric surgery);^12^ and -6.2 cmCSF (through low calorie diet).^13^ The efficacy in these studies is akin to what we have observed following treatment with exenatide for 3 months (-5.6 cmCSF).

Headache is a key disabling feature for idiopathic intracranial hypertension patients and listed in the top 10 priority areas for research by patient groups.^24^ Headache is also a key determinant for the significantly reduced quality of life in idiopathic intracranial hypertension.^20^ In this trial, whilst not powered to evaluate secondary outcome measures, there was a significant reduction in monthly headache days between baseline and 12 weeks amongst those patients taking exenatide (-7.7 days); those in the placebo arm did not have any significant improvement (-1.5 days) (Supplementary Table 4, Figure 4). A reduction in monthly headache of 1.5-2 days is regarded as meaningful in chronic migraine randomised controlled trials.^42-45^ Improvements in headache in idiopathic intracranial hypertension have been shown in other trials to occur due to reduction in ICP.^21^ Headache outcomes may show more benefit over a longer trial duration, as during the relatively short time horizon of this trial other factors may also have contributed to headache such as medication overuse and opiate use which occur in approximately one third of idiopathic intracranial hypertension patients.^42^

There was a significant improvement in visual acuity between the trial arms at 12 weeks (Supplementary Table 4, Figure 3). In this trial, the magnitude of change (5 letters) was equivalent to a line on the visual acuity chart. In other ocular diseases a change of > 5 letters would have > 90% probability of being a real change.^46^ Changes in visual acuity of 5 letters have been noted previously in a prospective crossover cohort study evaluating a weight loss intervention where there was a reduction of ICP by -6.2 cmCSF.^13^ It is likely that the improvement in visual acuity found here may be related to resolution of the hyperopic shift associated with raised ICP and papilloedema.

Weight loss has been shown to reduce ICP in idiopathic intracranial hypertension.^12,13^ In this study, there was no significant reduction in body weight at 12 weeks in the treatment arm, which implies that the reduction in ICP was highly likely to have been mediated by a reduction in CSF secretion, the mechanism demonstrated in pre-clinical studies, rather than through weight loss.^7^ Additionally, the significant reduction in ICP noted at the early time points of 2.5 hours and 24 hours would have been too premature to be modulated by weight loss. GLP-1 receptor agonists such as exenatide can increase satiety and promote weight loss, but this is generally in a context of a concurrent low calorie diet.^47^

Exenatide is widely used to treat type 2 diabetes and has an established long-term safety record (licensed in 2005).^48,49^ In this trial exenatide was safe and well-tolerated, with no withdrawals due to drug adverse effects. This is relevant as the drug predominantly used off-label for idiopathic intracranial hypertension, acetazolamide, can be poorly tolerated. In a previous randomised controlled trial 48% of patients withdrew over a three month period, due to adverse effects.^39^ Nausea is a known side effect of GLP-1 receptor agonists,^48,49^ and was predicted to occur in those idiopathic intracranial hypertension patients taking exenatide. In this trial 7 patients experienced nausea related to exenatide, but this settled over the first week. The nausea may, in part, be explained by the exenatide pharmacokinetic profile with high peak levels noted twice a day with twice daily Byetta dosing.^50^ Our pharmacokinetic data illustrated highest serum levels at 2.5 hours post-dose in keeping with the established literature on Byetta. Anti-drug antibodies were detected in two participants. The clinical relevance of this is not determined; however, anti-drug antibodies have not been shown to impact drug effects in diabetes.^33^ idiopathic intracranial hypertension is known to be a disease of insulin resistance,^51^ and therefore exenatide’s insulin sensitizing effects may have additional benefits in this population.

This is the first trial, to our knowledge, to utilise telemetric intracranial monitoring to measure ICP in idiopathic intracranial hypertension. Patient input into the trial design advocated for telemetric ICP monitoring over multiple lumbar punctures to facilitate data validity and reduce patient discomfort from multiple lumbar punctures. This has permitted detailed characterisation of ICP changes and the ability to measure ICP for prolonged periods over several weeks without further invasive procedures.^52^ This technology has led to the accurate demonstration of ICP lowering both with single dosing and with repeated dosing. The intracranial telemetric catheters were safe, with only one failing to register by 12 weeks.

This study has several limitations. As an early phase study, it was powered to the primary endpoints of ICP reduction with single and repeated dosing. Thus, the study was underpowered to detect differences in patient centred outcomes and the secondary clinical endpoints such as headache and visual field perimetry. Powering the study for those endpoints would have required a large increase in number of participants, however, telemetric ICP monitors would not be feasible in large trial cohort. No stratification was used at randomisation and groups were unmatched for baseline headache days and visual field perimetry. Additionally, patients with minimal visual field defects were not excluded as in prior studies thus making the study more clinically representative, but visual effects more difficult to detect due to the ceiling effect of the measurement.

The results of this study may have wide reaching implications for many other diseases of raised ICP as the drug does not target the specific pathogenesis of idiopathic intracranial hypertension, but CSF secretion. Given the tolerability of exenatide and its direct and early effect on ICP, it may be beneficial in other conditions such as hydrocephalus, traumatic brain injury, raised ICP in stroke and meningitis, and space flight associated neuro-ocular syndrome. In addition, GLP-1 receptor agonists have been shown to demonstrate neuro-protective properties and hence may have additional benefits in conditions such as traumatic brain injury.^53^

We had previously identified a novel pathway to modulate ICP in an animal model.^7^ This pre-clinical work has now been translated to a population of patients with raised ICP due to idiopathic intracranial hypertension. The data demonstrated the efficacy of exenatide to significantly lower ICP. A new efficacious therapy is a clear unmet patient need,^24^ and this is the first trial of a new drug treatment for idiopathic intracranial hypertension that significantly reduces ICP. The data presented here supports further evaluation of exenatide in a large randomized controlled trial, powered to evaluate clinically relevant outcome measures.

## Data Availability

All data produced in the present study are available upon reasonable request to the authors

## Abbreviations

ADA=: anti-drug antibodies;
A/G ratio=: andro-gynoid ratio;
BMI=: body mass index;
DEXA=: dual energy x-ray absorptiometry;
DPP-4=: dipeptidyl-peptidase 4;
GLP-1=: glucagon-like peptide 1;
HbA1C,=: glycated haemoglobin;
HCG=: human choroid gonadotrophin;
HIT-6=: headache impact test 6;
HOMA2-IR=: Homeostasis model assessment of insulin resistance;
HVF=: Humphrey visual field;
ICP=: intracranial pressure;
IIH=: idiopathic intracranial hypertension;
logMAR=: logarithm of the minimum angle of resolution,
LP=: lumbar puncture;
MCS=: mental component score;
MHD=: monthly headache days;
NRS=: numerical rating sale;
OCT=: optical coherence tomography;
PCS=: physical component score;
PIS=: patient information sheet;
PMD=: perimetric mean deviation;
PK=: pharmacokinetics;
RNFL=: retinal nerve fibre layer;
SF-36=: short form 36;
VA=: visual acuity,
VRS=: verbal rating scale.

## Competing interests

JLM was funded by the UK Ministry of Defence for the duration of the study. OG reports scientific consultancy fees from Invex therapeutics (2020).

AY reports receiving speaker fees from Teva, UK, outside the submitted work. KB works for UCB.

Professor Mollan reports other Invex Therapeutics, other Heidelberg engineering during the conduct of the study; other from Chugai-Roche Ltd, other from Janssen, other from Allergan, other from Santen, other from Roche, other from Neurodiem, outside the submitted work.

Professor Sinclair reports personal fees from Invex therapeutics in her role as Director with stock holdings, during the conduct of the study (since 28.06.2019); other from Allergan, Novartis, Cheisi and Amgen outside the submitted work.

All other authors declare no competing interests.

## Supplementary material

Supplementary material is available at *Brain* online.

---

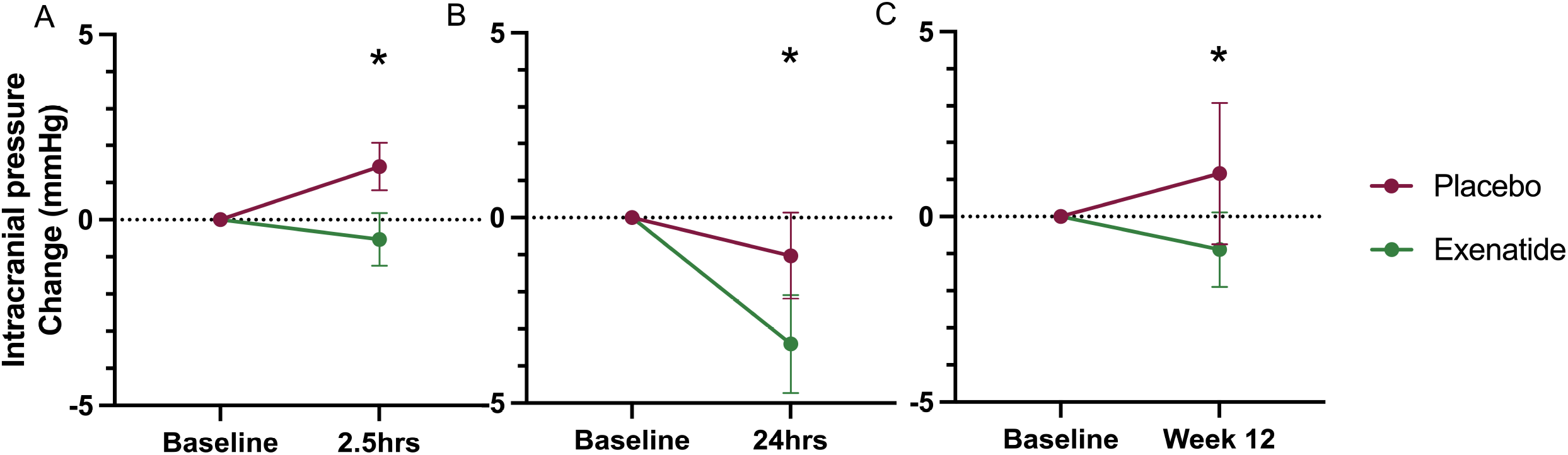

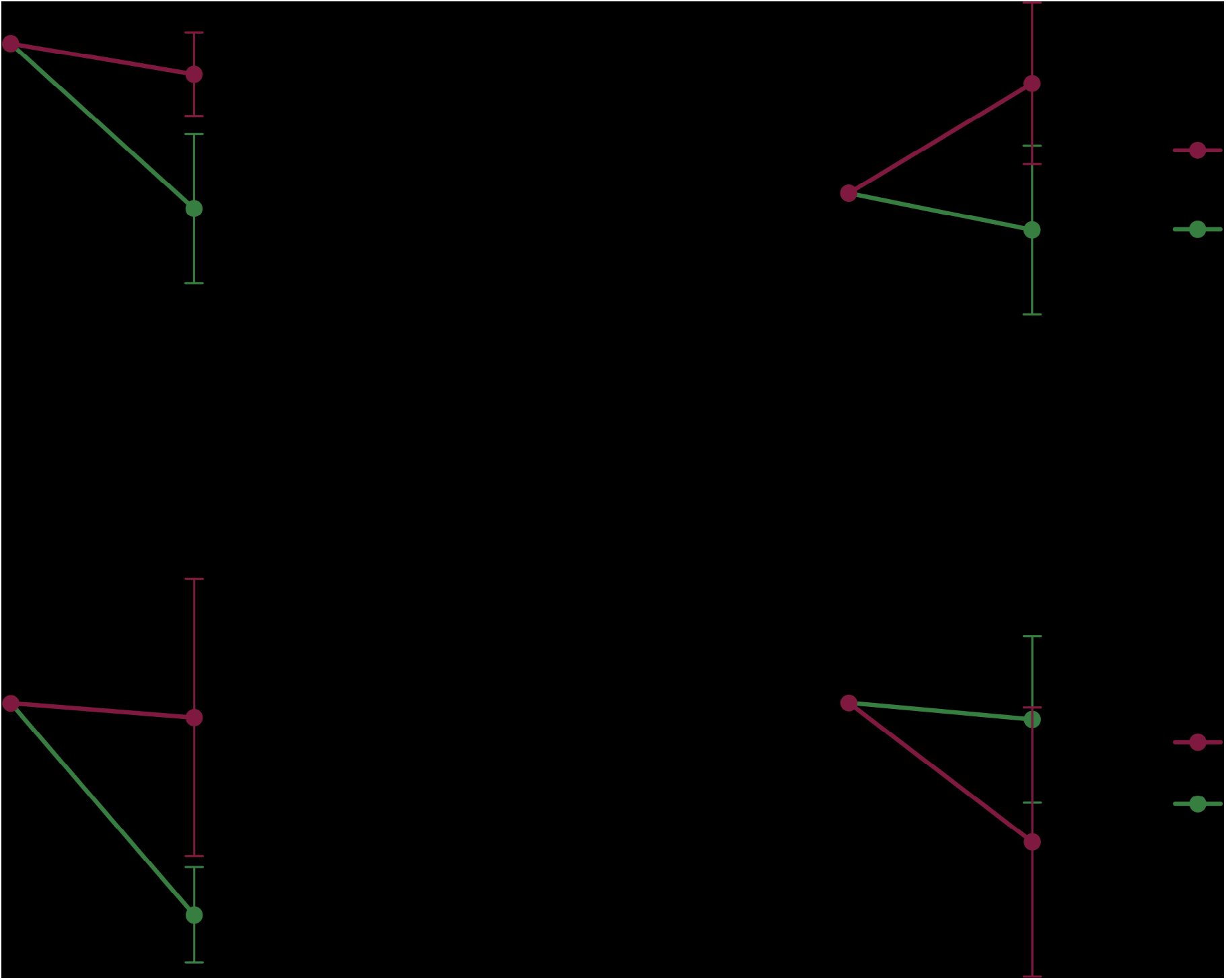

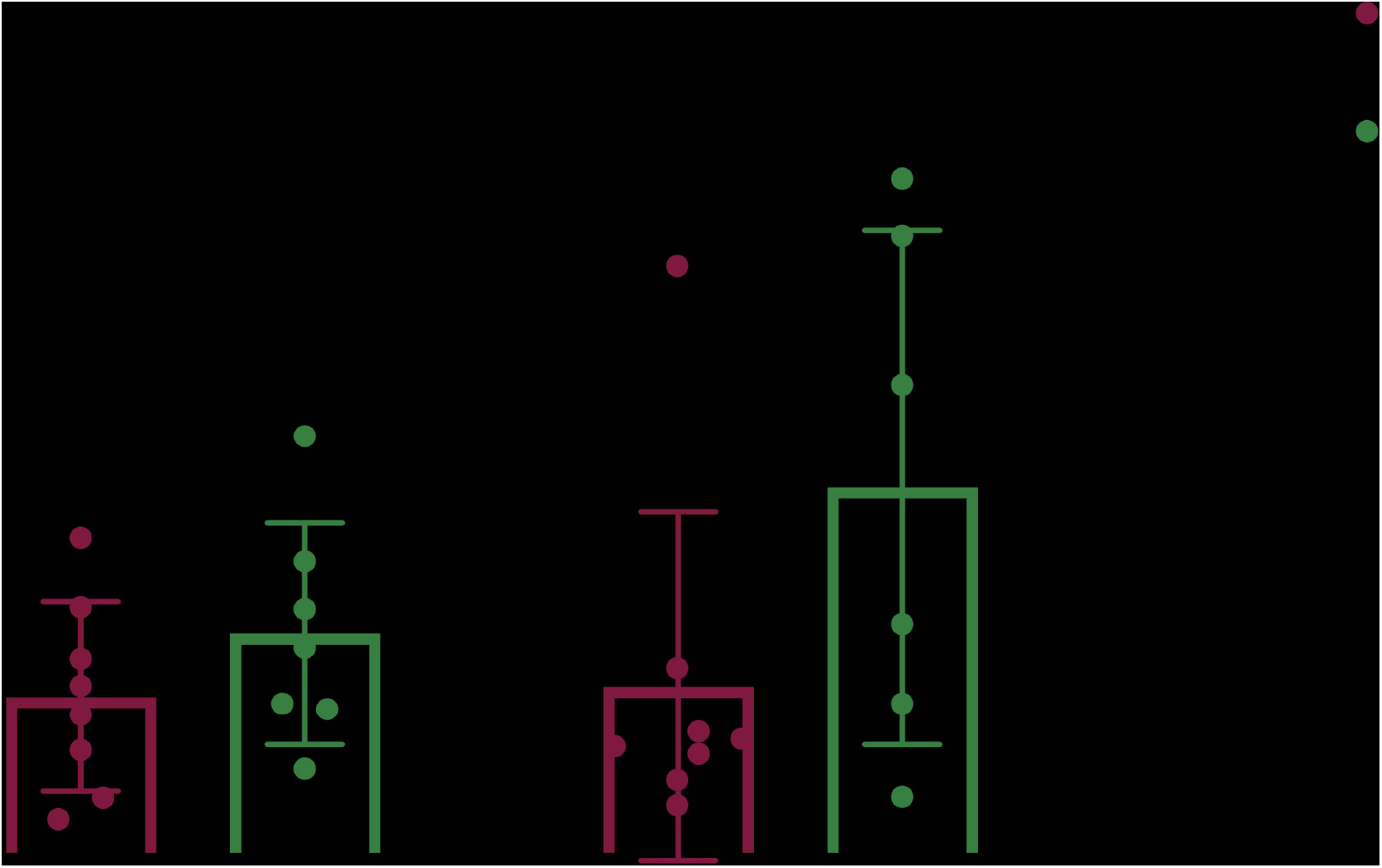

