## Supplemental tables for "The effect of GLP-1RA exenatide on Idiopathic Intracranial Hypertension: Randomised Clinical Trial"

Supplementary tables and figures.

| **Inclusion Criteria** |
|---|
| Female IIH patients aged between 18 and 60 years, diagnosed according to the modified Dandy criteria who have active disease (papilloedema [Frisen grade ≥ 1], significantly raised ICP > 25cm$H_2O$) and no evidence of venous sinus thrombosis (magnetic resonance imaging (MRI) or computerised tomography (CT) imaging and venography as noted at diagnosis). |
| Able to give informed consent. |
| **Exclusion Criteria** |
| Age less than 18 or older than 60 years. |
| Pregnant or trying to conceive. |
| Significant co-morbidity; such that in the opinion of the investigator it would not be in the participant's best interest to participate in the trial. |
| Addison's or Cushing's disease. |
| Functioning CSF shunt/stent or optic nerve sheath fenestration. |
| Currently using GLP-1 agonist or DPP-4 inhibitor. |
| Surgical contra-indication. |
| Concomitant therapy with acetazolamide, topiramate or diuretics (this can be discontinued 1 month prior to enrolment). |
| Inability to give informed consent e.g. due to cognitive impairment. |

**Supplementary Table 1 Inclusion and exclusion criteria.**

| **SCHEDULE OF EVENTS** |
|---|

| | Enrolment | Surgical implant | Baseline | | | | | | Treatment period | | |
|---|---|---|---|---|---|---|---|---|---|---|---|
| **Week** | -4 (minimum) | | 1 | | | | | | 2 | 8 | 12 |
| **Day** | | | 1 | | | | | 2 | 14 | 56 | 84 |
| **Hour** | | | pre-dose | 0 | 2.5 | 6 | 11 | 24 | | | |
| **Identification, trial and eligibility discussion, provision of PIS** | X | | | | | | | | | | |
| **Eligibility** | X | | | | | | | | | | |
| **Informed consent signature** | X | | | | | | | | | | |
| **Medical History** | X | | X | | | | | | | | |
| **HCG testing** | X | | X | | | | | | | | |
| **Provision of 4-week headache diary** | X | | | | | | | | | X | |
| **Physical Examination and clinical measurements** | X | | X | | | | | | | | X |
| **Insertion of ICP monitor** | | X | | | | | | | | | |
| **Randomisation** | | | X | | | | | | | | |
| **Telemetric pressure monitoring (ICP monitoring)** | | | X[2] | | X[3] | | | X[3] | | | X[3] |
| **LogMAR Visual Acuity** | | | X | | | | | | | | X |
| **Intraocular pressure** | | | X | | | | | | | | X |
| **Perimetric Mean Deviation (PMD – HVF)** | | | X | | | | | | | | X |
| **Optical Coherence Tomography RNFL** | | | X | | | | | | | | X |
| **Headache diary review** | | | X | | | | | | | | X |
| **BMI** | | | X | | | | | | | | X |
| **Blood pressure and heart rate** | x | x | x | | | | | | | | x |
| **Questionnaires: HIT-6 and SF-36** | | | X | | | | | | | | X |
| **Blood sampling – Biochemistry and ADA** | | | X | | | | | | X | | X |
| **Blood sampling PK** | | | X | X | X | X | X | X | | X | X |
| **DEXA** | | | X | | | | | | | | X |

**Supplementary Table 2 Schedule of events.**

| | All Mean (SD) | Exenatide Mean (SD) | Placebo Mean (SD) |
|---|---|---|---|
| **Number (n)** | 15 | 7 | 8 |
| **Age** | 28 (9) | 28 (13) | 28 (6) |
| **BMI (kg/m2)** | 38.1 (6.2) | 37.6 (7.9) | 38.6 (4.7) |
| **ICP (supine) mmHg** | 23.5 (3.9) | 22.3 (3.6) | 24.6 (4.1) |
| **ICP (LP position) cm CSF** | 32.2 (5.6) | 30.7 (6.7) | 33.5 (5.6) |
| **Frisen Grade (Worst eye)* median(IQR)** | 2 (1) | 2 (1) | 2.5 (1) |
| | **Median (IQR)** | **Median (IQR)** | **Median (IQR)** |
| **Duration of IIH at enrolment (months)** | **7 (35.5)** | **4 (15.5)** | **18.5 (55.3)** |
| **Time from surgery to baseline Time from surgery to baseline** | 10 (16.5)10 (16.5) | 4 (3.5) | 18.5 (19.5) |

**Supplementary Table 3 Baseline characteristics.**

| | Baseline mean (SD), n | 12 weeks mean (SD), n | Difference baseline to 12 weeks mean (SD); 95%CI, p | Difference between arms at 12 weeks mean (SE); 95%CI, p Hierarchical regression |
|---|---|---|---|---|
| **Monthly headache days** | | | | |
| **Exenatide** | 21.6 (5.2), n=7 | 13.9 (7.2), n=7 | -7.7 (9.2); (-16.3, 0.8), p=0.069 | 5.1 (3.9); (-2.4, 12.7), p=0.184 |
| **Placebo** | 10.3 (8.5), n=8 | 8.8 (8.0), n=8 | -1.5 (4.8); (-5.5, 2.5), p=0.404 | |
| **Monthly analgesic frequency** | | | | |
| **Exenatide** | 7.9 (4.5), n=7 | 7.0 (5.7), n=7 | -0.9 (5.2); (-5.7, 4.0), p=0.680 | 1.1 (2.5); (-3.8, 6.0), p=0.648 |
| **Placebo** | 3.4 (2.8), n=7 | 5.9 (5.2), n=7 | 2.4 (5.1); (-2.3, 7.1), p=0.254 | |
| **LogMar visual acuity** | | | | |
| **Exenatide** | 0.0 (0.05), n=7 | -0.1 (0.07), n=7 | -0.1 (0.04); (-0.1, 0.0), p=0.004 | -0.1 (0.05); (-0.2, -0.1), p=0.036 |
| **Placebo** | 0.0 (0.14), n=8 | 0.0 (0.12), n=8 | 0.0 (0.14); (-0.1, 0.1), p=0.921 | |
| **Perimetric mean deviation worst eye dB (HVF 24-2 sita standard)** | | | | |

| | | | | |
|---|---|---|---|---|
| **Exenatide** | -0.6 (1.0), n=7 | -1.0 (0.9), n=7 | -0.3 (1.1); (-1.4, 0.7), p=0.472 | 1.0 (0.8); (-0.5, 2.5), p=0.188 |
| **Placebo** | -2.7 (1.9), n=8 | -2.0 (1.6), n=8 | 0.7 (0.7); (0.1, 1.3), p=0.020 | |
| **Intraocular pressure** | | | | |
| **Exenatide** | 18.0 (2.0), n=7 | 16.9 (1.7), n=7 | -1.2 (2.8); (-3.8, 1.4), p=0.306 | -0.1 (1.1); (-2.3, 2.1), p=0.910 |
| **Placebo** | 16.7 (2.6), n=8 | 16.9 (1.9), n=7 | 0.5 (1.3); (-0.7, 1.7), p=0.375 | |
| **Optical Coherence Tomography RNFL worst eye (µm)** | | | | |
| **Exenatide** | 153 (58.9), n=6 | 132 (34.0), n=6 | -21.0 (28.8); (-51.2, 9.2), p=0.134 | -40.2 (47.2); (-133.0, 52.4), p=0.396 |
| **Placebo** | 183 (100.0), n=8 | 172 (114.0), n=8 | -10.8 (88.0); (-84.3, 62.8), p=0.740 | |
| **Quality of Life (SF-36) PCS summary** | | | | |
| **Exenatide** | 49.7 (20.3), n=7 | 53.8 (23.4), n=7 | 4.1 (7.4); (-2.7, 10.9), p=0.191 | -5.7 (9.5); (-24.3, 12.9), p=0.550 |
| **Placebo** | 57.8 (16.9), n=8 | 59.5 (11.8), n=8 | 1.7 (9.5); (-6.2, 9.6), p=0.632 | |
| **Quality of Life (SF-36) MCS summary** | | | | |
| **Exenatide** | 43.4 (23.3), n=7 | 44.8 (24.2), n=7 | 1.4 (8.3); (-7.3, 10.1), p=0.692 | -2.3 (10.3); (-22.5, 18.0), p=0.826 |
| **Placebo** | 46.6 (17.2), n=8 | 46.9 (10.5), n=8 | 0.5 (16.3); (-14.6, 15.5), p=0.940 | |
| **BMI (kg/m2** | | | | |
| **Exenatide** | 37.6 (7.9), n=7 | 37.5 (7.4), n=7 | -0.1 (0.8); (-0.8, 0.7), p=0.851 | -0.6 (3.3); (-7.0, 5.8), p=0.854 |
| **Placebo** | 38.6 (4.7), n=8 | 38.1 (4.9), n=8 | -0.5 (1.3); (-1.6, 0.6), p=0.336 | |
| **Mean arterial pressure (mmHg)** | | | | unpaired t-Test |
| **Exenatide** | 92.3 (10.8), n=7 | 85.8 (6.2), n=7 | -6.5 (12.9); (-18.4, 5.4), p=0.23 | -6.7 (3.6); (-1.1, 14.5), p=0.088 |
| **Placebo** | 89.6 (6.2), n=8 | 92.4 (5.2), n=8 | 2.8 (6.4); (-2.6, 8.1), p=0.26 | |

**Supplementary Table 4 Secondary and exploratory outcomes.**

| | **Baseline** | | | **12 weeks** | | |
|---|---|---|---|---|---|---|
| | **Exenatide** | **Placebo** | **chi-squared p** | **Exenatide** | **Placebo** | **chi-squared p** |
| **Headache severity (VRS 0-10) Category** | | | 0.133 | | | 0.084 |
| **Mild, n (%)** | 0 (0%) | 3 (38%) | | 0 (0%) | 4 (50%) | |
| **Moderate** | 6 (86%) | 5 (62%) | | 6 (86%) | 3 (38%) | |
| **Severe** | 1 (14%) | 0 (0%) | | 1 (14%) | 1 (13%) | |

### Supplementary Table 5 Headache severity.

| | Baseline | | | 12 weeks | | |
|---|---|---|---|---|---|---|
| | Exenatide | Placebo | chi-squared p | Exenatide | Placebo | chi-squared p |
| **Headache disability (HIT-6) Category** | | | | | | |
| Little-to-no impact, n (%) | 0 (0%) | 2 (25%) | 0.218 | 0 (0%) | 3 (38%) | 0.088 |
| Moderate | 0 (0%) | 1 (13%) | | 1 (14%) | 0 (0%) | |
| Severe | 5 (71%) | 2 (25%) | | 4 (57%) | 3 (38%) | |
| Substantial | 2 (29%) | 3 (38%) | | 0 (0%) | 2 (25%) | |
| Missing | | | | 2 (29%) | | |

### Supplementary Table 6. Headache disability.

| | Baseline mean (SD), n | 12 weeks mean (SD), n | Difference baseline to 12 weeks mean (SD); 95%CI, p | Difference between arms at 12 weeks mean (SE); 95%CI, p (Hierarchical regression) |
|---|---|---|---|---|
| **Creatinine (µmol/L)** | | | | |
| Exenatide | 67.6 (9.5), n=7 | 72.7 (3.5), n=7 | 5.1 (7.4); (-1.7, 12.0), p=0.117 | |
| Placebo | 66.4 (8.1), n=8 | 67.1 (9.7), n=8 | 0.8 (7.0); (-5.1, 6.6), p=0.770 | 5.6 (4.2); (-2.7, 13.9), p=0.186 |
| **Alanine transaminase (IU/L)** | | | | |
| Exenatide | 27.2 (13.9), n=6 | 24.9 (17.2), n=7 | -0.2 (7.9); (-8.4, 8.1), p=0.961 | |
| Placebo | 21.3 (11.7), n=8 | 16.8 (5.4), n=8 | -4.4 (7.3); (-10.5, 1.7), p=0.134 | 8.0 (6.5); (-4.7, 20.7), p=0.218 |
| **High density lipoprotein (mmol/L)** | | | | |
| Exenatide | 1.3 (0.4), n=7 | 1.2 (0.2), n=7 | 0.0 (0.3); (-0.3, 0.2), p=0.710 | |
| Placebo | 1.5 (0.2), n=8 | 1.5 (0.3), n=8 | 0.0 (0.2); (-0.2, 0.2), p=0.866 | -0.2 (0.1); (-0.5, 0.0), p=0.088 |
| **Cholesterol (mmol/L)** | | | | |
| Exenatide | 4.5 (0.8), n=7 | 4.7 (1.0), n=7 | 0.2 (0.6); (-0.4, 0.7), p=0.472 | |
| Placebo | 4.8 (1.0), n=8 | 4.7 (1.4), n=8 | -0.1 (0.7); (-0.6, 0.5), p=0.715 | 0.0 (0.6); (-1.1, 1.1), p=0.964 |
| **Triglycerides (mmol/L)** | | | | |

| | | | | |
|---|---|---|---|---|
| Exenatide | 1.3 (0.6), n=7 | 1.3 (0.5), n=7 | 0.1 (0.5); (-0.4, 0.6), p=0.680 | 0.2 (0.2); (-0.3, 0.6), p=0.488 |
| Placebo | 1.1 (0.2), n=8 | 1.2 (0.4), n=7 | 0.0 (0.4); (-0.3, 0.4), p=0.782 | |
| **HbA1C (mmol/mol)** | | | | |
| Exenatide | 35.4 (2.7), n=5 | 36.3 (2.9), n=7 | 1.0 (1.4); (-0.8, 2.8), p=0.189 | 1.8 (1.6); (-1.4, 5.0), p=0.278 |
| Placebo | 35.0 (3.9), n=6 | 34.5 (3.0), n=8 | 0.7 (2.7); (-2.2, 3.5), p=0.576 | |

**Supplementary Table 7 Blood test results.**

| | **Number of events** | **Arm** | | **Severity** | **Relatedness** | **Description** |
|---|---|---|---|---|---|---|
| | | Exenatide n | Placebo n | | | |
| **Adverse Events** | | | | | | |
| **Nausea** | 3 | 3 | 0 | Moderate | Related | Nausea requiring treatment on baseline visit |
| **Nausea** | 4 | 4 | 0 | Mild | Related | Mild transient nausea |
| **Minor wound infection** | 3 | 1 | 2 | Mild | Unrelated | Participant continued in trial |
| **Post-Operative facial swelling** | 1 | 0 | 1 | Mild | Unrelated | Participant continued in trial |
| **Serious Adverse Events** | | | | | | |
| **Thyrotoxicosis** | 1 | 0 | 1 | Moderate | Unrelated | Participant continued in trial |
| **Withdrawals** | | | | | | |
| **Withdrawals** | 1 | 1 | 0 | | | Pre-randomisation |

**Supplementary Table 8 Adverse events.**

| Timepoint | Day 0 20 µg Exenatide pg/ml (mean (SD)) n=7 | Day 14 10 µg Exenatide BD pg/ml (mean (SD)) n=4 | Day 84 10 µg Exenatide BD pg/ml (mean (SD)) n=4 |
|---|---|---|---|

| | | | |
|---|---|---|---|
| 0 | 44.4 (12.0) | 81.3 (54.7) | 91.6 (61.8) |
| 2.5 | 575.4 (501.6) | 380.7 (180.4) | 205.4 (83.2 |
| 6 | 180.9 (84.9) | | |
| 11 | 67.3 (22.5) | | |
| 22 | 44.6 (16.8) | | |
| 24 | 335.8 (106.5) | | |

**Supplementary Table 9 Exenatide serum concentration.**

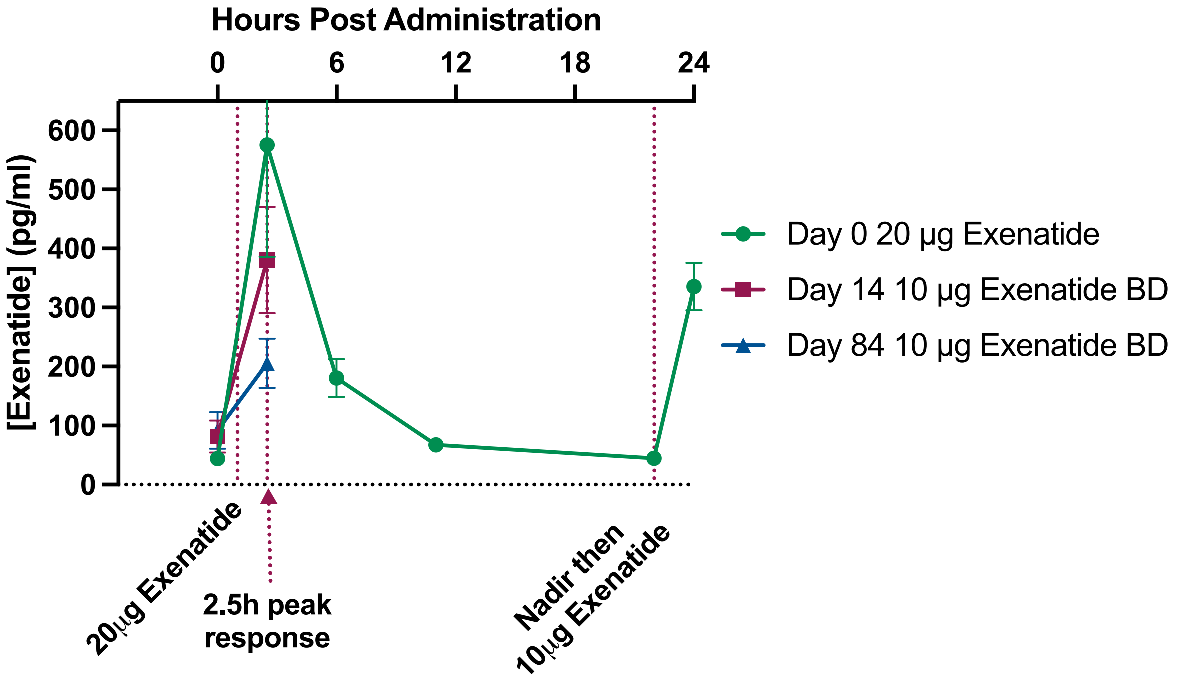

**Supplementary Figure 1 Exenatide serum concentration (mean and SEM).** Serum exenatide levels were measured after single bolus administration of exenatide 20µg on day 0 and following 10µg dosing on day 14 and 84.

| | Baseline mean (SD), n | 12 weeks mean (SD), n | Difference baseline to 12 weeks T-test, mean (SD); 95%CI, p | Difference between arms at 12 weeks T-test, mean (SE); 95%CI, p |
|---|---|---|---|---|
| **Insulin** | | | | |
| Exenatide | 98.0 (49.4), 6 | 163.2 (114.6), 6 | 59.6 (91.75); (-36.7, 155.9) p=0.17 | 88.6 (51.2); -23.0, 200.3) p=0.11 |
| Placebo | 70.0 (42.3), 8 | 74.5 (77.8), 8 | 4.6 (59.1); (-44.9, 54.0) p=0.83 | |
| **Glucose** | | | | |
| Exenatide | 3.2 (0.7), 5 | 3.6 (0.5), 5 | 0.5 (1.0); (-0.8, 1.7) p=0.36 | |

| | | | | |
|---|---|---|---|---|
| Placebo | 3.2 (0.3), 8 | 3.4 (0.4), 8 | 0.1 (0.2); (-0.05, 0.3) p=0.13 | 0.26 (0.26); (-0.3, 0.8) p=0.34 |

**Supplementary Table 10 Insulin and glucose serum concentration.**

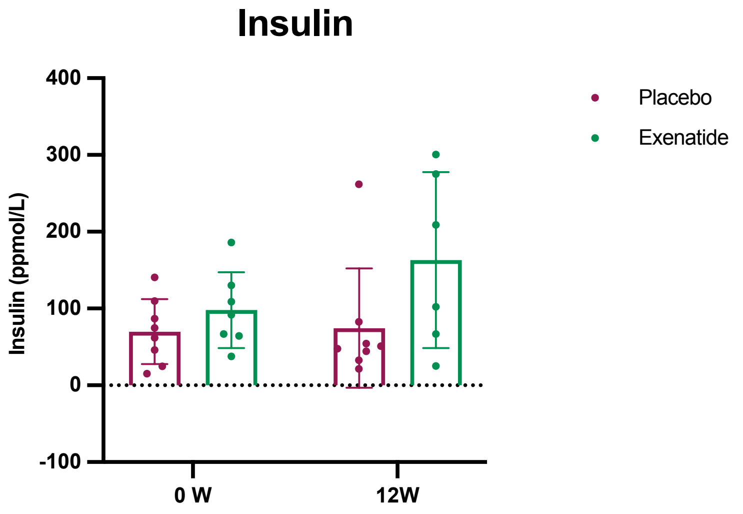

**Supplementary Figure 2 Insulin and glucose serum concentration**. Blood insulin and glucose concentrations were measured at baseline and 12 weeks.

| Time | Baseline ICP (mmHg) mean (SD) | At time point ICP (mmHg) mean (SD) | Difference baseline to time point mean (SD); 95%CI, p | Difference between arms at time point mean (SE); 95%CI, p (Hierarchical regression) |
|---|---|---|---|---|
| **0-30 mins** | | | | |
| Exenatide | 22.3 (3.6) | 23.3 (4.1) | 1.0 (1.3); (-0.2, 2.2), p=0.089 | -2.3 (2.1); (-6.3, 1.8), p=0.274 |
| Placebo | 24.6 (4.1) | 25.4 (4.0) | 1.6 (2.0); (-.03, 3.4), p=0.080 | |
| **30-60 mins** | | | | |
| Exenatide | 22.3 (3.6) | 23.7 (4.9) | 1.4 (2.1); (-0.5, 3.3), p=0.125 | -3.4 (2.1); (-7.5, 0.7), p=0.106 |
| Placebo | 24.6 (4.1) | 26.3 (4.8) | 2.5 (2.8); (-0.1, 5.1), p=0.055 | |
| **60-90 mins** | | | | |
| Exenatide | 22.3 (3.6) | 22.6 (4.7) | 0.3 (1.9); (-1.4, 2.1), p=0.637 | -3.4 (2.1); (-7.5, 0.7), p=0.102 |
| Placebo | 24.6 (4.1) | 25.3 (2.8) | 1.5 (2.6); (-0.9, 3.9), p=0.174 | |
| **90-120 mins** | | | | |
| Exenatide | 22.3 (3.6) | 22.0 (3.8) | -0.4 (2.8); (-3.3, 2.6), p=0.769 | -4.6 (2.1); (-8.8, -0.5), p=0.028 |
| Placebo | 24.6 (4.1) | 25.7 (2.4) | 1.8 (2.7); (-0.6, 4.3), p=0.116 | |
| **120-150 mins** | | | | |
| Exenatide | 22.3 (3.6) | 21.8 (3.4) | -0.5 (1.9); (-2.3, 1.2), p=0.485 | -4.2 (2.1); (-8.3, -0.2), p=0.042 |
| Placebo | 24.6 (4.1) | 26.0 (3.4) | 1.4 (1.8); (-0.1, 2.9), p=0.060 | |

**Supplementary table 11 ICP monitoring, 2.5-hour time course.** Due to the nature of hierarchical analysis, there is slight variation in values compared to those of the primary outcomes, however data included at timepoints is identical.

| Time | At time point ICP (mmHg) mean (SD) | Difference between arms at time point mean (SE); 95%CI, p (Hierarchical regression) |
|---|---|---|
| **Midnight (baseline) – 01:00** | | |
| Exenatide | 16.1 (3.5) | -2.5 (2.3); (-7.1, 2.1), p=0.284 |
| Placebo | 18.2 (5.7) | |
| **01:00 – 02:00** | | |
| Exenatide | 14.7 (3.2) | -4.0 (2.3); (-8.4, 0.5), p=0.082 |
| Placebo | 18.7 (4.0) | |
| **02:00 – 03:00** | | |
| Exenatide | 15.2 (3.8) | -1.9 (2.3); (-6.4, 2.6), p=0.406 |
| Placebo | 17.1 (3.9) | |
| **03:00 – 04:00** | | |
| Exenatide | 15.4 (2.2) | -2.9 (2.3); (-7.4, 1.6), p=0.206 |
| Placebo | 18.3 (4.4) | |
| **04:00 – 05:00** | | |
| Exenatide | 14.5 (4.8) | -5.6 (2.3); (-10.1, -1.0), p=0.018 |
| Placebo | 20.7 (4.1) | |
| **05:00 – 06:00** | | |
| Exenatide | 16.1 (3.2) | -4.2 (2.5); (-9.1, 0.6), p=0.086 |
| Placebo | 20.1 (3.1) | |
| **06:00 – 07:00** | | |
| Exenatide | 16.2 (6.0) | -5.0 (2.5); (-9.9, -0.2), p=0.040 |
| Placebo | 22.1 (4.1) | |

**Supplementary Table 12 Overnight ICP monitoring.**

| | Baseline mean (SD), n | 12 weeks mean (SD), n | Difference baseline to 12 weeks T-test, mean (SD); 95%CI, p | Difference between arms at 12 weeks T-test, mean (SE); 95%CI, p |
|---|---|---|---|---|
| **Fat % total** | | | | |
| Exenatide | 51.4 (3.1), n=7 | 51.2 (3.3), n=7 | -0.17 (1.1); (-1.2, 0.9), p=0.70 | 0.50 (1.8); (-3.4, 4.4), p=0.78 |
| Placebo | 51.9 (3.9), n=8 | 50.7 (3.6), n=8 | -1.2 (1.6); (-2.6, 0.08), p=0.06 | |
| **Fat mass** | | | | |
| Exenatide | 51.0 (16.5), n=7 | 51.2 (16.8), n=7 | 0.15 (0.14); (-1.1, 1.4), p=0.78 | 2.5 (7.0); (-12.7, 17.7), p=0.73 |
| Placebo | 50.9 (11.1), n=8 | 48.6 (10.1), n=8 | -2.3 (3.2); (-5.0, 0.4), p=0.086 | |

| | | | | |
|---|---|---|---|---|
| **Lean mass** | | | | |
| Exenatide | 47.0 (10.0), n=7 | 47.4 (10.1), n=7 | 0.4 (1.0); (-0.5, 1.4), p=0.31 | 0.8 (4.2); (-8.2, 9.7), p=0.86 |
| Placebo | 46.3 (5.4), n=8 | 46.6 (5.7), n=8 | 0.29 (1.5); (-0.9, 1.5), p=0.60 | |
| **DEXA A/G ratio** | | | | |
| Exenatide | 1.1 (0.08), n=7 | 1.1 (0.08), n=7 | -0.009 (0.04); (-0.04, 0.03) p=0.57 | 0.05 (0.04); (-0.05, 0.1), p=0.28 |
| Placebo | 1.1 (0.08), n=8 | 1.1 (0.09), n=8 | 0.001 (0.03); (-0.02, 0.02), p=0.90 | |

**Supplementary Table 13 DEXA body composition analysis.**
