## Supplementary material for "The effect of GLP-1RA exenatide on Idiopathic Intracranial Hypertension: Randomised Clinical Trial": Table 1

|  | **ICP at baseline mean (SD)** | **ICP at time point mean (SD)** | **Difference in ICP baseline to time point mean (SD); 95%CI, p** | **Difference in ICP between arms at time point mean (SE); 95%CI, p** |
| --- | --- | --- | --- | --- |
| **ICP 2.5 hours (mmHg)** | | | | |
| **Exenatide (n=7)** | 22.3 (3.6) | 21.8 (3.4) | -0.5 (1.9); (-2.3, 1.2), p=0.485 | -4.2 (2.1); (-8.4, 0.0), p=0.048 |
| **Placebo (n=8)** | 24.6 (4.1) | 26.0 (3.4) | 1.4 (1.8); (-0.1, 2.9), p=0.060 |  |
| **ICP 24 hours (mmHg)** | | | | |
| **Exenatide (n=7)** | 22.3 (3.6) | 18.9 (5.3) | -3.4 (3.5); (-6.6, -0.2), p=0.042 | -4.7 (2.1); (-8.8, -0.5), p=0.030 |
| **Placebo (n=8)** | 24.6 (4.1) | 23.5 (4.5) | -1.0 (3.3); (-3.8, 1.7), p=0.406 |  |
| **ICP 12 weeks (mmHg)** | | | | |
| **Exenatide (n=7)** | 22.3 (3.6) | 21.4 (4.0) | -0.9 (2.7); (-3.3, 1.6), p=0.410 | -4.1 (2.2); (-8.4, 0.1), p=0.058 |
| **Placebo (n=7)** | 24.6 (4.1) | 26.0 (4.4) | 1.2 (5.1); (-3.5, 5.8), p=0.565 |  |

**Table 1.** Primary outcome measures. ICP, intracranial pressure.
